# Overexpression of long non coding RNA OR3A4 is associated with altered p38 signalling, morphological and phenotypic changes, and reduced immunogenicity in Barrett’s Oesophagus

**DOI:** 10.1101/2021.05.29.21258052

**Authors:** Tom Nieto, Yash Sinha, Qin Qin Zhuang, Mathew Coleman, Joanne Stockton, Celina Whalley, Rahul Hejmadi, Mark Dilworth, Agata Stoldona, Valerie Pestinger, Olga Tucker, Andrew D Beggs

## Abstract

**Background:** Barrett’s Oesophagus (BO) presents a particular pathological dilemma, in that patients who have no dysplasia within their BO experience a small but significant risk of malignant progression each year. Screening programmes have attempted to reduce the mortality from BO associated oesophageal adenocarcinoma but cannot predict which BO patients will progress to invasive malignancy. We have previously identified the long non coding RNA, *OR3A4*, is differentially hypomethylated in progressive BO. We aimed to understand its role in BO pathogenicity

**Methods:** The stable BO cell line CP-A, as well as the oesophageal adenocarcinoma cells line OE-33 was transfected with a lentiviral OR3A4 over-expression vector, and underwent high resolution microscopy, immunofluorescence, RT-qPCR, RNA sequencing, and targeted drug screening with the p38-MAPK inhibitor domipramod to understand the effects of OR3A4 expression on progression. We then compared progressive vs. non-progressive BO samples using quantitative multi-fluorophore (Vectra) immunohistochemistry.

**Results:** Over-expression of OR3A4 in CP-A lines resulted in a hyperproliferative, dysplastic cellular phenotype, with strong over-expression of MAPK and anti-apoptotic pathways at the RNA and protein level, which was sensitive to the p38-MAPK inhibitor domipramod. Vectra immunohistochemistry demonstrated that progressive BO had reduced visibility associated with a reduction in CD8+ T-cells and CD68+ macrophages and reduced CD4+ T-cells in the stomal compartment.

**Conclusion:** The overexpression of OR3A4, which we have previously shown is associated with progressive BO leads to a proliferative dysplastic cellular phenotype associated with increased, reversible MAPK signalling and loss of immune visibility.

## BACKGROUND

Long non coding RNA (LncRNA), defined as non-translated RNA that are greater than 200 nucleotides in length, play a key role in regulation of gene expression (1). Dysregulation of lncRNAs have been associated with numerous human cancers (2, 3). For example, The HOX antisense intergenic RNA (HOTAIR) has been found to be upregulated in breast cancer and especially in metastatic breast cancer (4) leading to metastasis via binding and remodelling chromatin at the polycomb repressive complex 2 (PRC2). Other lncRNAs identified as associated with cancer include Metastasis associated lung adenocarcinoma transcript 1 (MALAT1) (5) in non-small cell lung cancer and HULC in hepatocellular cancer, colorectal liver metastases, gastric cancer, pancreatic cancer and osteosarcoma (6). lncRNAs can also act as tumour suppressors as is the case with maternally expressed gene 3 (MEG3). Over-expression of MEG3 appears to activate p53 tumour suppressing pathways and increase p53 levels in vitro (7). In HeLa (cervical cancer), MCF-7 (breast cancer) and H4 (neuroglioma) cell lines, over-expression of MEG3 inhibits growth (8). The breadth of LncRNA dysregulation in different types of cancer, and the mechanistic role in tumorigenesis in different tissues is not yet clear. Their role as prognostic indicators and therapeutic targets in different cancers of unmet need is increasing.

Barrett’s oesophagus (BO) represents the probable pre-malignant lesion in oesophageal adenocarcinoma (OADC), with approximately 2-3% of the population estimated to have BO (9). Dysplasia within BO is an independent risk factor for malignancy, however the majority of BO is non-dysplastic (NDBO) and it is difficult to identify patients with NDBO who will progress to OADC (10). We have previously shown that patients who progress from NDBO to OADC have relative hypomethylation within a region proximal to the LncRNA OR3A4 (11) that drives increased OR3A4 expression.

Initially, the long non-coding RNA OR3A4 was thought to have no functional relevance, but has recently been identified as an LncRNA. Guo et al (12) investigated the biological relevance of OR3A4 in gastric cancer, demonstrating that OR3A4 expression is upregulated in metastatic gastric cancer tissues compared to normal gastric and gastric cancer tissues. OR3A4 expression in 130 paired tumour/normal gastric cancer tissue samples correlated with lymph node metastasis, depth of tumour invasion and distal metastasis as well as being increased in the plasma of patients with gastric cancer.

In vitro models showed increased proliferation and cell migration/invasion of SGC7901 (gastric cancer) cells transfected with an OR3A4 over expressing vector(12), and decreased proliferation of NCI-N87 cells (known to over-express OR3A4) when siRNA knockdown of OR3A4 was performed (an effect replicated in mouse models). Target enrichment in this model demonstrated that several potential molecular mechanisms for OR3A4 increasing tumourigenicity.

Liu et al (13) also reported that OR3A4 is upregulated in breast cancer tissues and cell lines in comparison to adjacent normal tissue and non-transformed cell lines, correlating with lymph node status, differentiation, ER status and TNM grading as well as being independently predictive of survival. On siRNA mediated knockout of OR3A4 in two breast cancer lines, MDA-MB-468 and MCF-7, a growth suppressive effect was seen.

Overall, and in association with our findings in BO, the LncRNA OR3A4 is over-expressed in multiple tumour types. Although OR3A4 is selected for enhanced growth in multiple cancers, its role in BO, NDBO and OC, and the molecular mechanisms involved, are not yet known. Therefore, we aimed to investigate the functional effect of OR3A4 over-expression in BO and OADC cell lines in vitro by creating stable cell lines which over-expressed OR3A4 in order to study its downstream effects in this tissue type.

## RESULTS

### Cell transfections

In order to understand the functional relevance of OR3A4 hypomethylation and resultant over-expression in the progression of BO to OADC, OR3A4 was over-expressed to simulate the effect of OR3A4 promoter hypomethylation in target cell lines. The NDBO cell line CP-A, and OADC line OE-33, were selected as experimental lines and were transduced with either an empty lentiviral vector or one expressing OR3A4 cDNA.

Following viral transduction and puromycin selection4 stable cell lines were generated; CP-A PIPZ control, CP-A PIPZ OR3A4, OE-33 PIPZ Control and OE-33 PIPZ OR3A4.

Once established and validated in stable culture, the transfected control and experimental lines were first analysed by real time quantitative PCR (RT-qPCR) to confirm over-expression of OR3A4. CP-A PIPZ OR3A4 cells exhibited a 13.7 (95%CI +/- 2) log-fold increase in expression of OR3A4 compared to CP-A normal controls, whereas the CP-A PIPZ control cells showed a 0.26 (95%CI +/- 0.01) log fold increase in OR3A4 expression. Similarly, the OE-33 PIPZ OR3A4 cell line exhibits a 13.8 (95%CI +/- 2.0) log-fold increase in expression of OR3A4 when compared to the OE-33 normal controls, whereas the OE-33 PIPZ controls show a 0.58 (95%CI +/- 0.51) log-fold increase.

### OR3A4 over-expression causes morphological changes in Barrett’s oesophagus cells

Next, we observed the OR3A4 stable cell lines by standard light microscopy to investigate any macroscopic morphological changes that were associated with OR3A4 over-expression. When viewed at high power at 50% confluence, CP-A cells are relatively homogenous with a spindle like appearance (Figure 1A). The empty PIPZ vector did not significantly change the cell morphology (Figure 1B). However, when OR3A4 was over-expressed in the CP-A line, the cells became larger and less homogenous in appearance (median perimeter 87.5 vs. 57.2, p<0.001). The nuclei of OR3A4 over-expressing cells also appeared enlarged compared to the controls as determined by image cytometry (median nuclei diameter 58.4 vs. 28.53, p<0.001), with enlarged nucleoli (Figure 1C). At 100% confluence, both the un-transfected cells and control transfected cells begin to lose the spindle like morphology, but remain relatively homogenous with little variation between individual cells forming a “cobblestone” monolayer (Figure 1D and E). The OR3A4 over-expressing cells in contrast, display increased heterogeneity with variation in size and morphology forming a poorly organised monolayer with reduced cell-cell contacts (Figure 1F).

**Figure 1.**
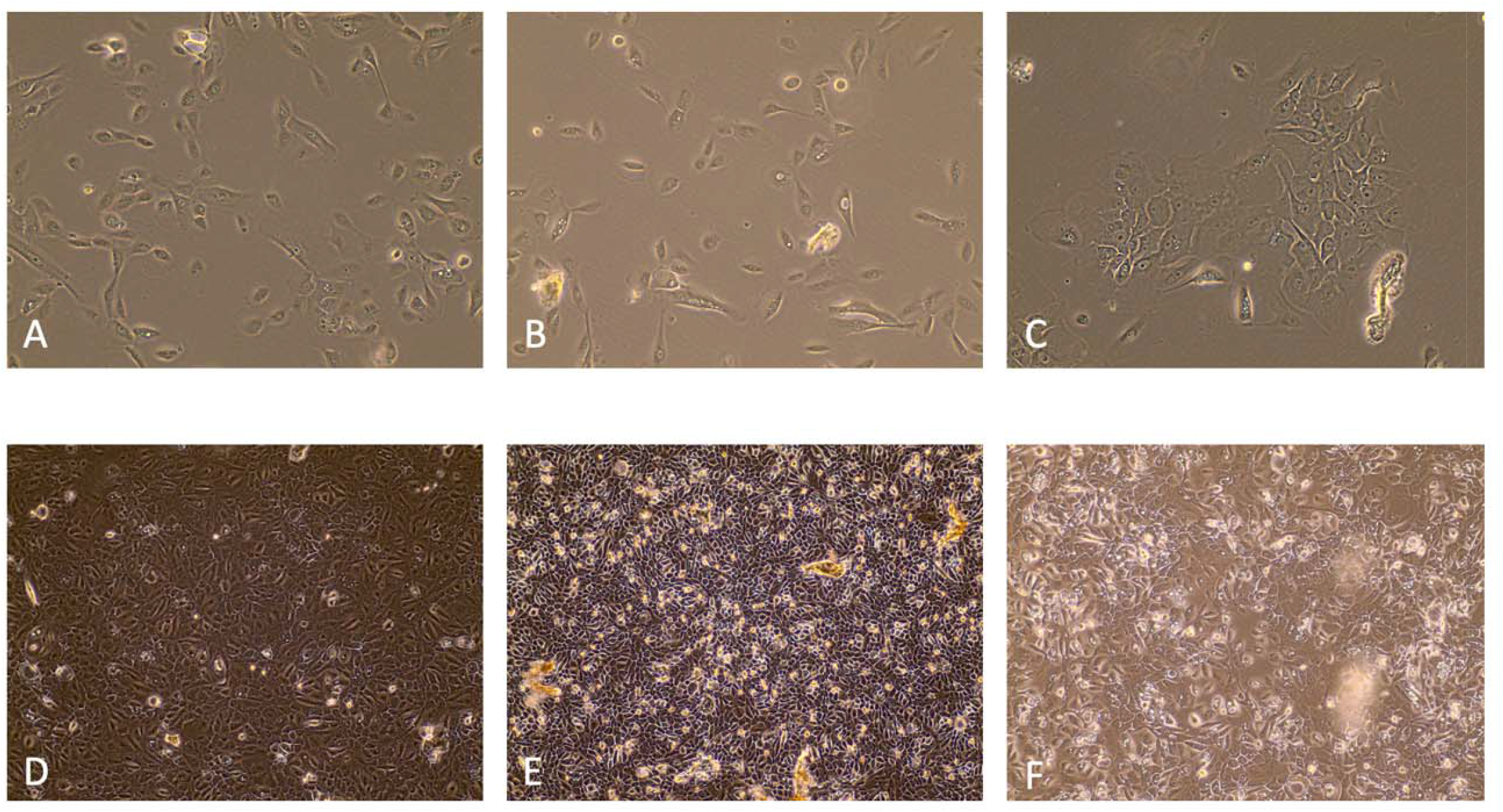
Light microscopy of CP-A line with and without transfection A: CPA parental cell line (40X), B: CPA PIPZ control (40X), C: CPA PIPZ OR3A4 over-expressing cells (40X), D: CPA parental cell line at 100% Confluence (10X), E: CPA PIPZ Control at 100% Confluence (10X), F: CPA PIPZ OR3A4 Over-Expressing Cells at 100% Confluence (10X).

When viewed under light microscopy, the morphological differences between the OE-33 control and experimental cell lines were less pronounced than between the BO cell lines. The cells demonstrated an epithelioid morphology and the OR3A4 over-expressing OE-33 cells (Figure 2C) maintained a similar nuclear to cytoplasmic ratio (median 0.25 vs. 0.29. p=0.342) and outline when compared with both PIPZ control transfected (Figure 2B) and non-transfected OE-33 cells (Figure 2A). The nuclei also retain a typical granular appearance across the experimental and control lines.

**Figure 2.**
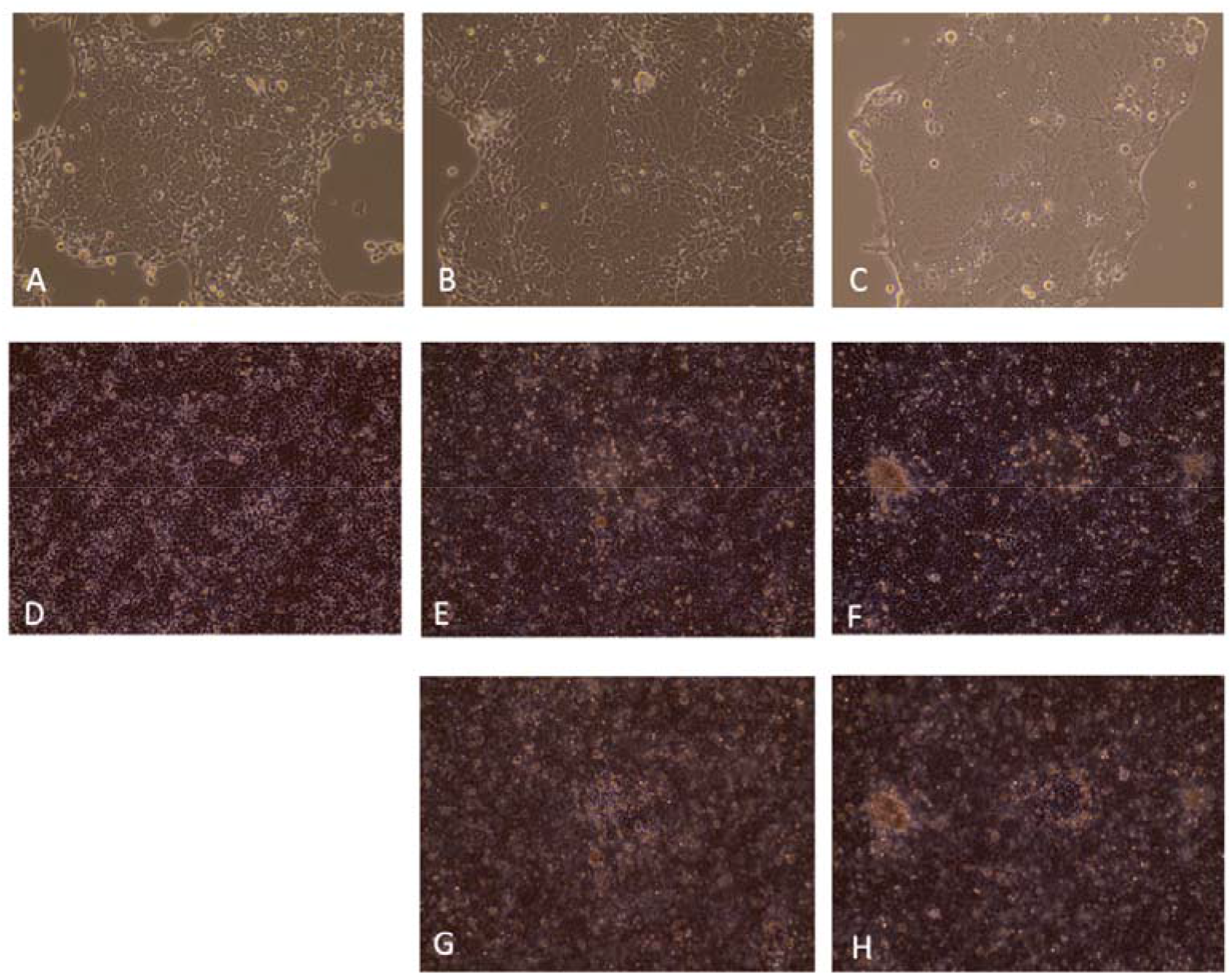
Light microscopy of OE-33 line with and without transfection A: OE-33 parental cell line (40X), B: OE-33 PIPZ control (40X), C: OE-33 PIPZ OR3A4 over-expressing cells (40X), D: OE-33 parental cell line at 100% Confluence (10X), E: OE-33 PIPZ Control at 100% Confluence (10X), F: OE-33 PIPZ OR3A4 Over-Expressing Cells at 100% Confluence (10X). G & H: change in focus of E & F respectively to demonstrate dome formation.

**Figure 3.**
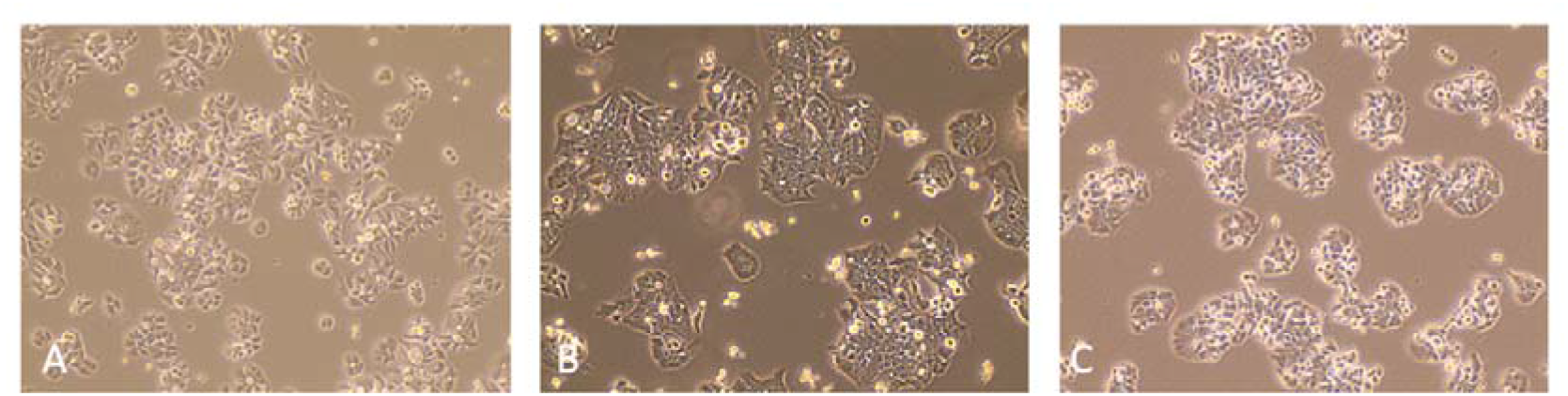
A: Light microscopy of OE19 line with and without transfection. OE-19 parental cell line (40X), B: OE-19 PIPZ control (40X), C: OE-19 PIPZ OR3A4 over-expressing cells (40X). Transfected cells did not grow to confluence.

To further characterise and highlight morphological differences (immunofluorescence (IF) was performed in CP-A and OE-33 cell lines using both control transfected and OR3A4 over-expressing lines. The images enhanced the morphological differences between CP-A and CP-A PIPZ OR3A4 with an increased nuclear:cytoplasmic ratio clearly evident and a dysplastic appearance of the cells with irregular cell wall outline and nuclei. The CP-A control cells display a homogenous, spindle like appearance (Figure 4A) whereas the OR3A4 over-expressing line appear to lose this morphology with increased cellular heterogeneity (Figure 4B).

**Figure 4.**
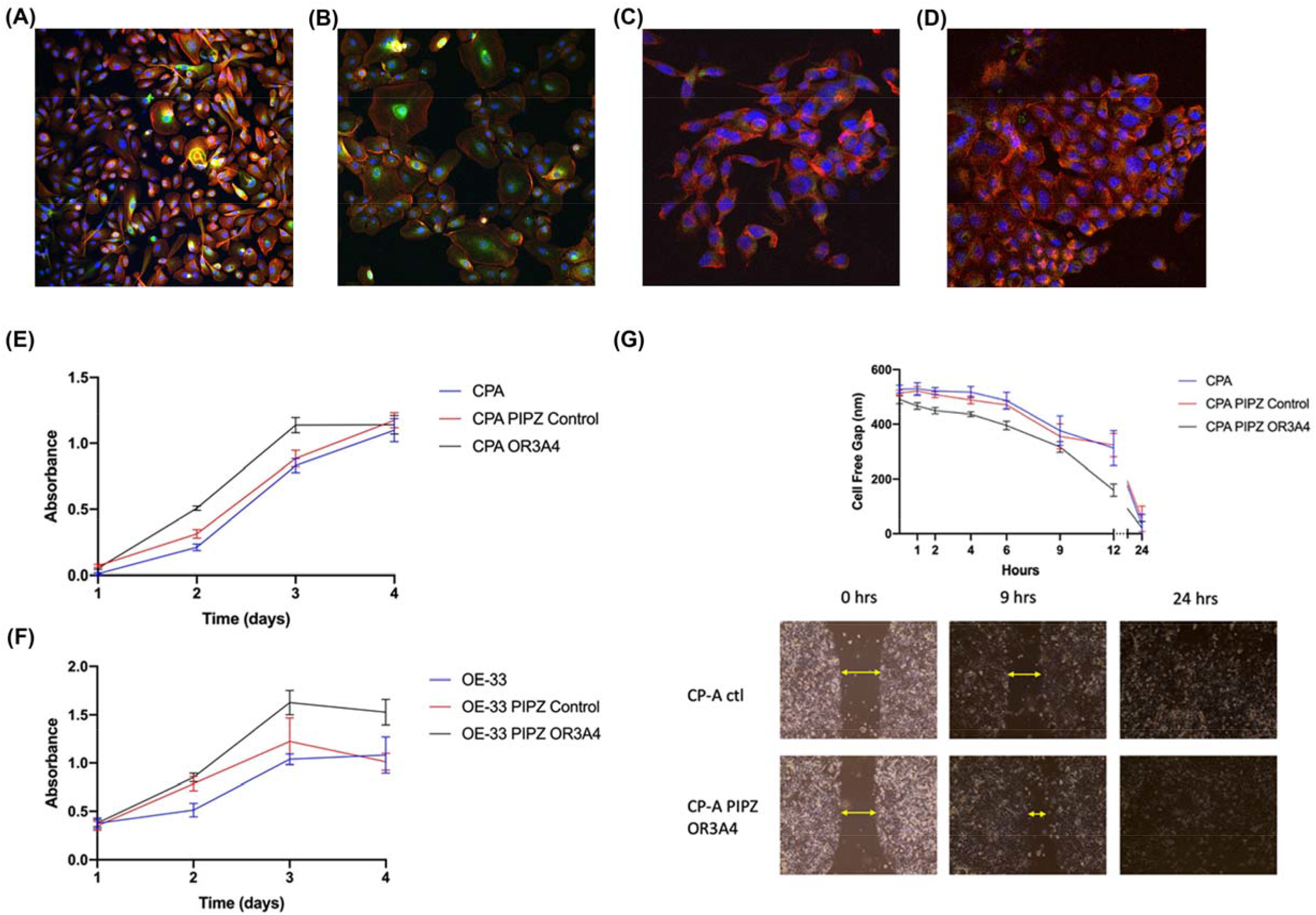
IF image of control and transfected cells. A- Cells stained with DAPI nuclear counter stain (blue), Phalloidin (red) and Caspase 3 (yellow). A- IF Image of CPA Control Cells x10. B- IF Image of CPA OR3A4 Over-Expressing Cells x10. C- IF Image of OE-33 Control Transfected Cell Line x10. D- IF image of OE-33 Cell Line Over-Expressing OR3A4 x10. E- MTS Assay: CP-A, CP-A PIPZ Control and CP-A PIPZ OR3A4. Absorbance at 490nm readings taken 4 hours after MTS reagent added F- MTS assay: OE-33, OE-33 PIPZ control AND OE-33 PIPZ OR3A4. Absorbance readings taken 4 hours after MTS reagent added. G- Scratch Wound Healing Assay. Cell free gap for CP-A control and CP-A PIPZ OR3A4 over closing time. The cells over-expressing OR3A4 closed the cell free gap and reached confluence more rapidly than the control lines.

**Figure 5:**
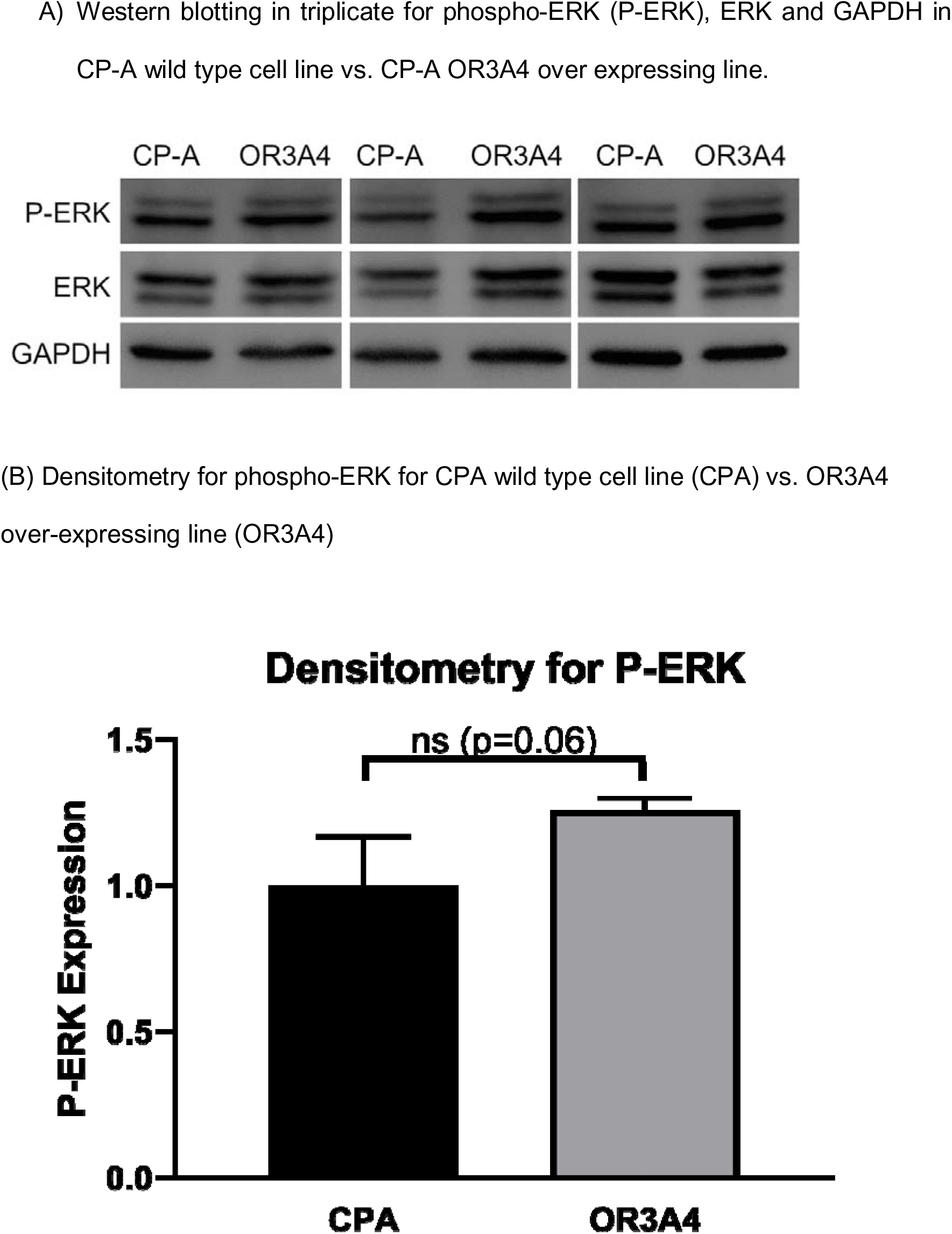

As seen under light microscopy, the morphology of the OE-33 cell lines was not appreciably different when enhanced by IF imaging. The control transfected (Figure 4C) and OR3A4 over-expressing (Figure 4D) cell lines both retained their homogenous epithelioid appearance and did not show changes in caspase 3 staining.

The changes observed might be expected to be associated with other gross phenotype alterations, particularly viability, migration and invasion so we next investigated these phenomena in the transfected lines.

### Increase in OR3A4 levels causes an increase in viability, migration and invasion in transfected cells

To assess the effect of OR3A4 expression on cell viability and growth rates, an MTS assay was performed in triplicate. When CP-A, CP-A PIPZ Control and CP-A PIPZ OR3A4 were analysed it demonstrated increased cell proliferation in the over-expressing cell lines compared to the control lines. Absorbance in the OR3A4 over-expressing line plateaued at 72 hours, whilst the control lines plateaued at 96 hours (Figure 4E). This indicates that the over-expressing cell lines reached confluence more rapidly than the controls.

Similar results were found in the OADC line OE-33 with OE-33 PIPZ OR3A4 cells reaching confluence more rapidly because of increased density (Figure 4F). The difference in cell proliferation and migration was assessed using a scratch wound healing assay using the protocol described in the materials and methods section. This assay assesses how rapidly cells migrate across and close a defined cell-free gap on a culture plate. Over a period of 24 hours, both CP-A and OE-33 cells which over-expressed OR3A4 closed the cell free gap more rapidly than both the transfected and normal control cell lines (p=0.0018). When the control cell lines were compared there was no significant difference between them.

### Increased OR3A4 levels lead to changes in MAP kinase signalling and apoptosis

To begin to investigate the mechanisms involved in the altered morphology, proliferation and migration of OR3A4 over-expressing cells we carried out RNA sequencing of the transfected lines vs. their controls. This was because lncRNAs regulate transcription and alternate transcripts and OR3A4 overexpression has previously been demonstrated to affect gene expression(12). The top 20 differentially expressed genes are presented in Table 1. Overall, there were 495 genes significantly under-expressed (p < 0.05 cut-off) by the control cell line compared to the experimental line and 618 were over expressed (p < 0.05 cut-off). The two most significantly upregulated genes were *PRKRIP1* (apoptosis) and *PRELID3A* (apoptosis) with increased fold change of 28.04 (p = 0.0002) and 12.89 (p = 0.0004) respectively. When we examined the expression of *PDLIM2*, which had previously (12) been shown to be downregulated in OR3A4 overexpression, we found a non-significant difference (LSMean(Control) = 0.66 vs. LSMean(OR3A4) = 0.16, p=0.08).

**Table 1.** Top 20 Differentially Expressed Genes of CPA Control Vs CPA OR3A4 Over-Expressing Cell Lines on RNA Sequencing

| Gene ID | Total counts | P-value<br>(Control vs.<br>OR3A4) | FDR step up<br>(Control vs.<br>OR3A4) | Fold change<br>(Control vs.<br>OR3A4) |
| --- | --- | --- | --- | --- |
| *OR3A4P* | 5.71E+02 | 4.72E-08 | 4.27E-03 | -17170.12 |
| *PRKRIP1* | 3.25E+00 | 2.24E-04 | 1.00E+00 | 28.04 |
| *SLMO1* | 3.49E+00 | 4.15E-04 | 1.00E+00 | 12.89 |
| *NCOA7* | 1.26E+00 | 4.16E-04 | 1.00E+00 | -34.69 |
| *SMCO4* | 8.61E-01 | 5.98E-04 | 1.00E+00 | -23.37 |
| *PCMTD2* | 6.96E+00 | 6.14E-04 | 1.00E+00 | 9.80 |
| *TXNL4A* | 7.63E-01 | 7.16E-04 | 1.00E+00 | -20.93 |
| *UBLCP1* | 1.54E+00 | 7.18E-04 | 1.00E+00 | 16.35 |
| *SETD5-*<br>*AS1* | 9.80E-01 | 7.93E-04 | 1.00E+00 | -20.98 |
| *FBXO36* | 8.52E-01 | 8.07E-04 | 1.00E+00 | -18.93 |
| *MRPL30* | 1.00E+00 | 8.28E-04 | 1.00E+00 | -20.78 |
| *CRYZL1* | 2.41E+00 | 8.57E-04 | 1.00E+00 | 23.03 |
| *ATRIP* | 6.88E-01 | 8.73E-04 | 1.00E+00 | -18.21 |
| *TSEN2* | 1.39E+00 | 9.59E-04 | 1.00E+00 | -17.08 |
| *GSR* | 8.41E-01 | 1.01E-03 | 1.00E+00 | -22.97 |
| *FAM86C1* | 6.98E-01 | 1.32E-03 | 1.00E+00 | -18.09 |
| *HMCES* | 5.79E-01 | 1.32E-03 | 1.00E+00 | -15.37 |
| *CEP41* | 5.98E-01 | 1.36E-03 | 1.00E+00 | -15.88 |
| *ATXN1* | 6.49E-01 | 1.45E-03 | 1.00E+00 | -14.45 |

To better understand the context of the differentially expressed genes discovered by total RNA sequencing, we undertook KEGG pathway based analysis using the’ database for annotation, visualisation and integrated discovery’ (DAVID) (14). The results, detailed in Table 2, revealed enrichment for altered regulation of apoptosis and MAPK pathways in cells overexpressing OR3A4

**Table 2:**
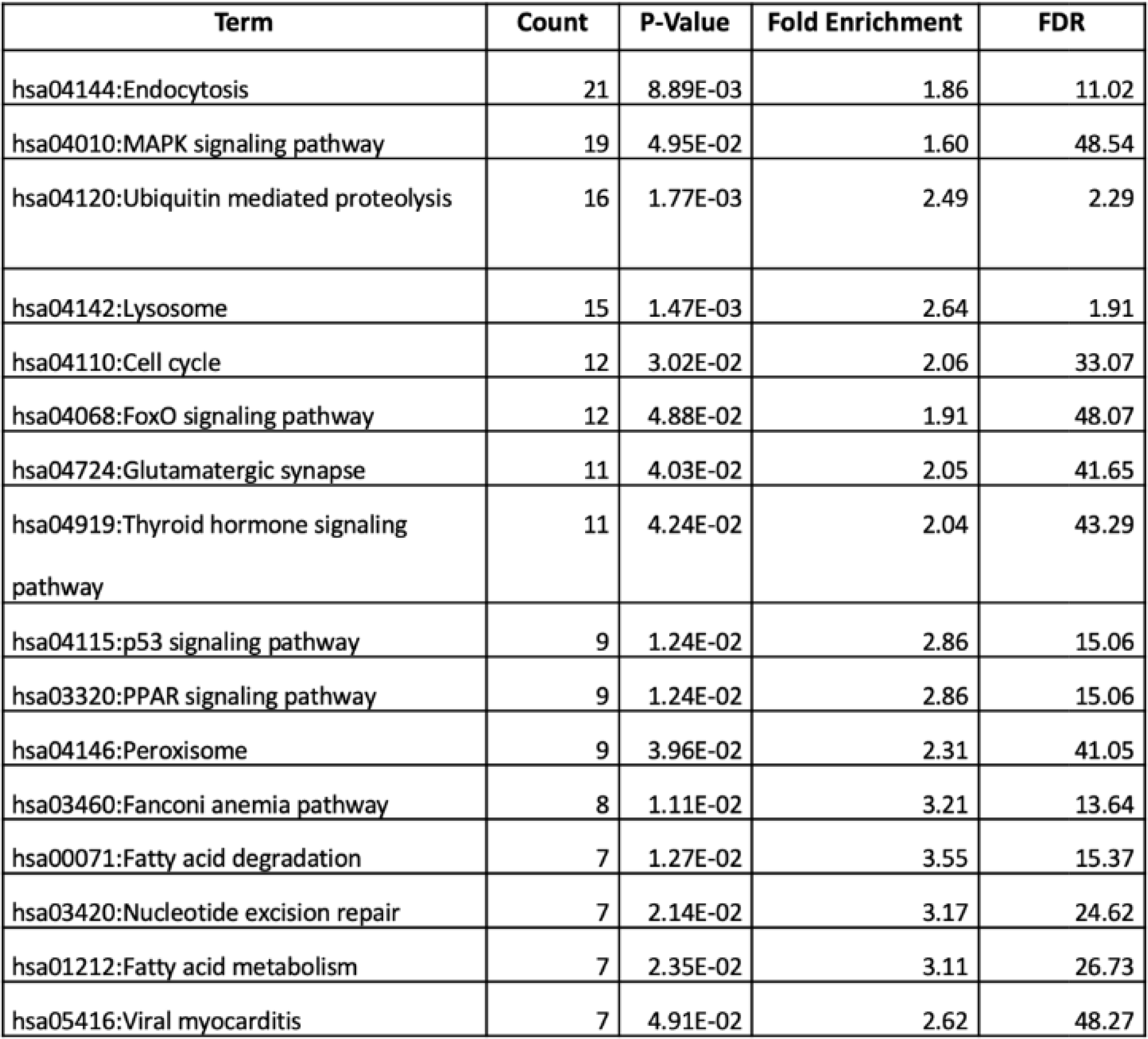
KEGG Enrichment Analysis

| Term | Count | P-Value | Fold Enrichment | FDR |
| --- | --- | --- | --- | --- |
| hsa04144:Endocytosis | 21 | 8.89E-03 | 1.86 | 11.02 |
| hsa04010:MAPK signaling pathway | 19 | 4.95E-02 | 1.60 | 48.54 |
| hsa04120:Ubiquitin mediated proteolysis | 16 | 1.77E-03 | 2.49 | 2.29 |
| hsa04142:Lysosome | 15 | 1.47E-03 | 2.64 | 1.91 |
| hsa04110:Cell cycle | 12 | 3.02E-02 | 2.06 | 33.07 |
| hsa04068:FoxO signaling pathway | 12 | 4.88E-02 | 1.91 | 48.07 |
| hsa04724:Glutamatergic synapse | 11 | 4.03E-02 | 2.05 | 41.65 |
| hsa04919:Thyroid hormone signaling pathway | 11 | 4.24E-02 | 2.04 | 43.29 |
| hsa04115:p53 signaling pathway | 9 | 1.24E-02 | 2.86 | 15.06 |
| hsa03320:PPAR signaling pathway | 9 | 1.24E-02 | 2.86 | 15.06 |
| hsa04146:Peroxisome | 9 | 3.96E-02 | 2.31 | 41.05 |
| hsa03460:Fanconi anemia pathway | 8 | 1.11E-02 | 3.21 | 13.64 |
| hsa00071:Fatty acid degradation | 7 | 1.27E-02 | 3.55 | 15.37 |
| hsa03420:Nucleotide excision repair | 7 | 2.14E-02 | 3.17 | 24.62 |
| hsa01212:Fatty acid metabolism | 7 | 2.35E-02 | 3.11 | 26.73 |
| hsa05416:Viral myocarditis | 7 | 4.91E-02 | 2.62 | 48.27 |

### Validation of over-expressed pathways reveals signalling changes in apoptosis

We found in the RNA-seq experiments that apoptosis was a differentially expressed pathway, with PRELID3A being one of the most significantly differentially expressed genes, being strongly over-expressed. PRELID3A exists in a complex with TRIAP1 which interacts with TP53 to regulate apoptosis. We also found that the MAPK pathway was over-expressed in the RNA-seq data which was of particular interest as MAPK pathways were of particular interest as these were enriched on pathway analysis in our previous whole genome methylation analysis of BO tissue samples which subsequently progressed to OADC (11).

Expression of PRELID3A and TRIAP1 (TP53 regulation of apoptosis 1) were analysed by RT-qPCR in CP-A, CP-A PIPZ Control, CPA PIPZ OR3A4, OE-33, OE-33 PIPZ Control and OE-33 PIPZ OR3A4 cell lines. Surprisingly, this showed no significant increase in expression of TRIAP1 or PRELID3A in the control or experimental cell lines using this method, suggesting that the RNA-seq result may have been a false positive. However, analyses of other transcripts within the human apoptosis RT-qPCR pathway array did reveal differences following OR3A4 expression. When OR3A4 was over expressed, there was an increase in DIABLO (31.38 log2 fold p=0.01), CASP14 (Caspase 14, 5.48 log2 fold, p=0.03) and DEDD2 (2.00 log2 fold, p=0.02). In the OR3A4 over-expressing cells lymphotoxin alpha (LT-α) had a 51.22 log2 fold downregulation (p<0.001) in comparison to the control cells. Combined, the dysregulated genes would supress apoptosis enhancing cell survival.

We then looked at changes in expression in MAPK pathways between control CP-A cells and OR3A4 over-expressing CP-A cells. Consistent with the RNA-seq pathway analysis there were significant changes (p <0.05) in regulation of the MAPK pathway between the two cell lines with increased expression of MAP2K3 (19.7 log2 fold), MAPK15 (11.2 log fold) and decreased expression of DUSP4 (7.48 log2 fold) expression., which in combination would cause upregulation of MAPK signalling.

We then investigated the downstream protein changes (Figure 6) seen in the MAPK pathway. Interestingly, although not quite reaching statistical significance, we did observe a 1.35-fold increase in phospho-ERK, a 2.5-fold increase in phospho-p38, and a 1.6-fold decrease in phospho-C-jun levels in OR3A4 overexpressing cells, suggesting differential regulation of MAPK pathways.

**Figure 6:**
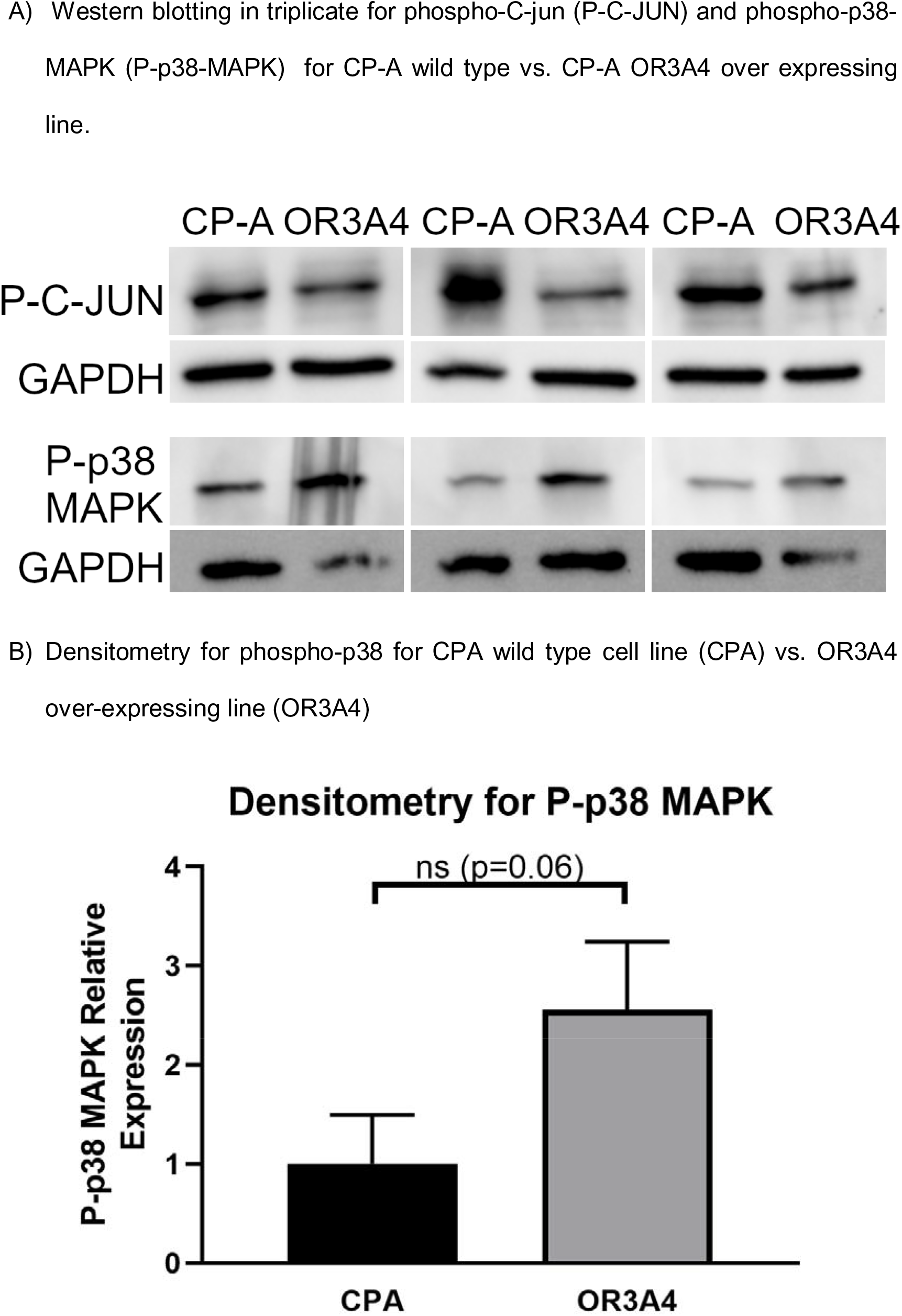
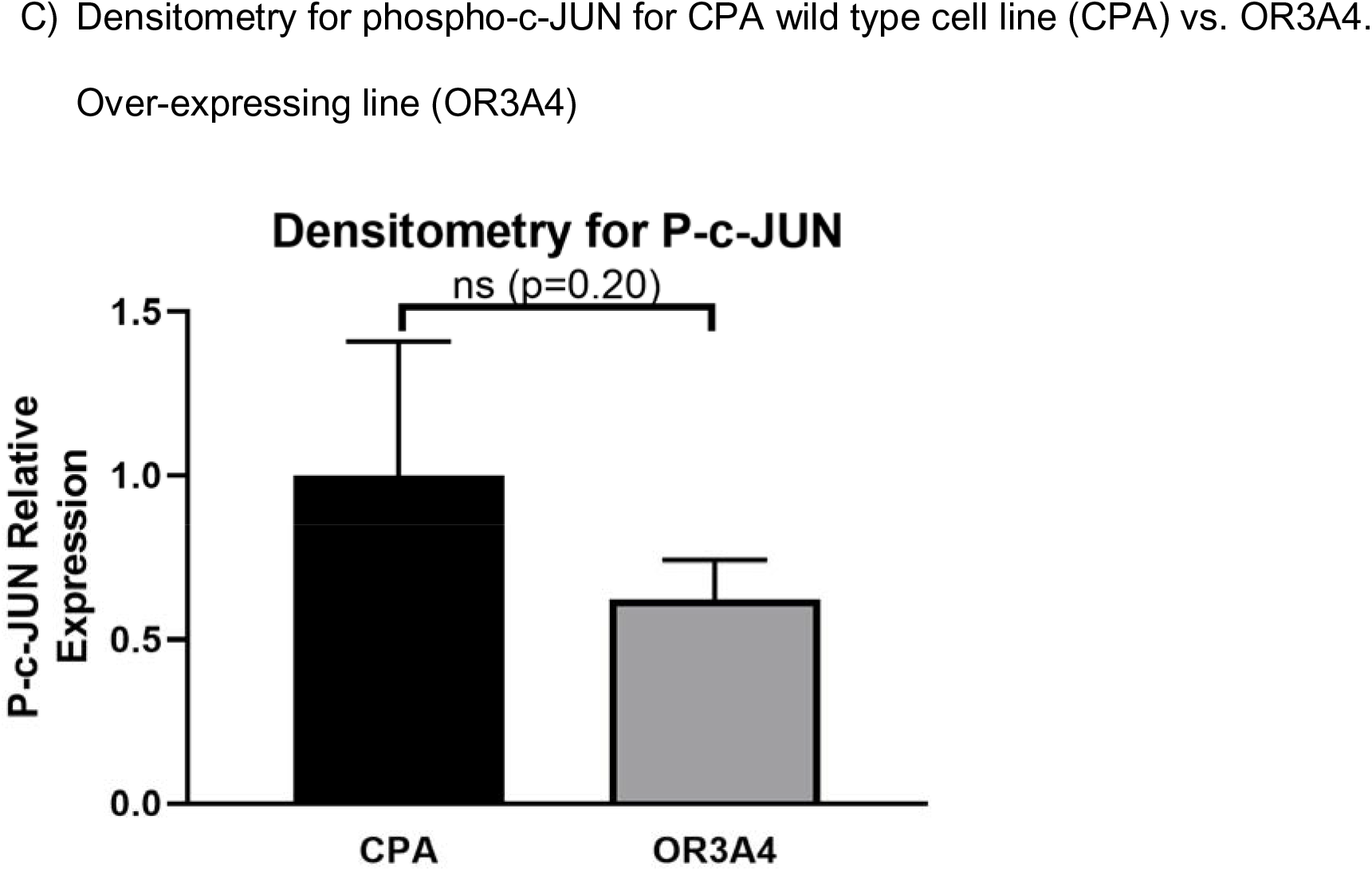

Overall, the transcriptional analyses identified OR3A4-dependent changes in apoptosis and MAPK signalling that could contribute to the morphological and phenotypic changes described above. Such changes could lead to specific ‘dependencies’ that could be exploited for therapeutic applications.

### Barrett’s oesophagus cells are sensitive to pharmacological inhibition of p38 signalling

To further investigate the functional relevance of the altered p38 MAPK pathway, and a potential pharmacological target for high risk BO, CP-A cells were treated with the p38 MAPK inhibitor Doramapimod (BIRB 796), which has entered clinical trials for the treatment of rheumatoid arthritis (15). CP-A, CP-A transfected control and CP-A OR3A4 cells were cultured and treated with increasing concentrations of Doramapimod from 0 to 10 μM. Their viability was then assessed with the MTS assay over the course of 5 days.

This revealed that OR3A4 over-expressing cells required higher concentrations of doramapimod to reduce viability compared to control cell lines, suggesting that OR3A4 increases MAPK signalling requiring increased blockade in order to supress growth.

### The immune landscape differs between progressor vs. non-progressor Barrett’s Oesophagus

Because of the known effects of p38 signalling on the immune landscape (16), and the relative non-visibility of Barrett’s oesophagus to the immune system, we hypothesised that the changes we had seen in p38 signalling in OR3A4 over-expressing cells (which are more likely to progress to dysplasia) could affect the immune context of Barrett’s. Sections of NDBO tissue from samples pre-dating any dysplastic change were taken from 12 BO progressor patients and from 9 non-progressor patients and stained using immunohistochemistry with a panel of antibodies including CD4, CD8, CC20, CD68 and FOXP3 (Tables 3-7). All sections were reported as non-dysplastic on histological review. These were scanned using the Vectra automated quantitative pathology imaging system and analysed with inForm software (Figure 7A). The software algorithm was trained to differentiate between the stroma, epithelium and background as shown in Figure 7B. The software was also trained to identify each individual cell subtype by the staining as shown in Figure 7C. Once the algorithm had been trained, images were batch processed and cells were identified and counted in the regions of interest using the InForm software.

**Figure 7:**
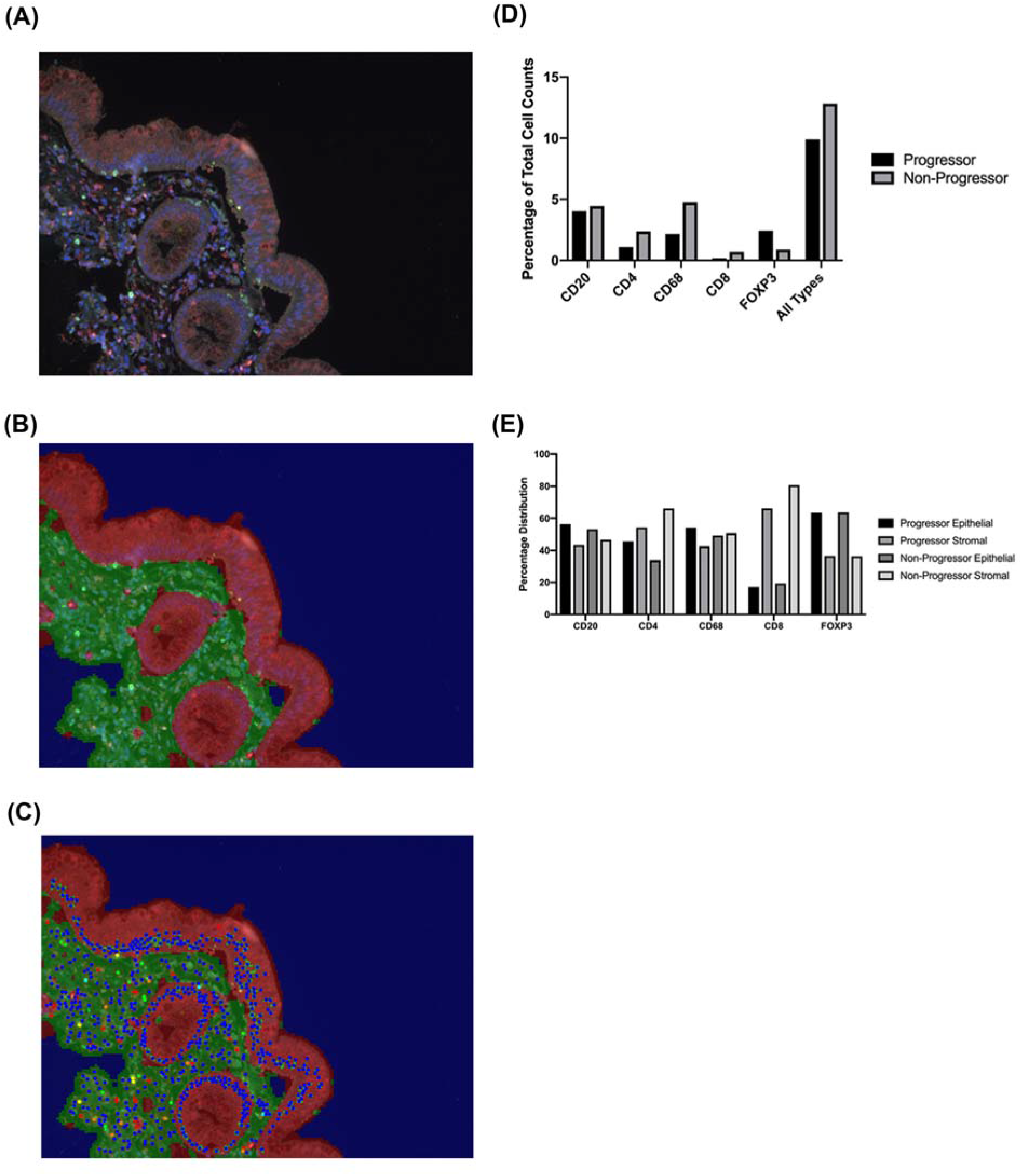
A - Image of NDBO tissue section following multiplex IHC staining taken with Vectra Imaging System. This image has the background fluorescence subtracted and leaves only the cellular staining of interest. This allows the InForm software to identify and differentiate between different cell types. The software algorithm was initially trained by manually selecting each individual cell type. Each cell was stained with a different OPAL dye corresponding to the light emission wavelength. CD68 690nm Red, FOXP3 650nm orange, CD20 620nm yellow, CD8 570nm green, CD4 520nm Cyan, DAPI nuclear Counterstain 460nm Blue. B - InForm Software tissue segmentation - The same tissue section is then segmented according to histological areas of interest. The epithelium is highlighted in red, stroma in green, background in blue. Areas are first highlighted manually, and the algorithm trained until it is capable of autonomously segmenting previously unseen images. This required manually segmenting multiple images to ensure accuracy of the final image processing. C - InForm Software Image with tissue segmentation and Cellular Identification. This image is the final result of the automated image processing. This allows the software to count the number and type of cells and give their distribution between the stroma and cytoplasm. D - Immune Cell Infiltration as a Percentage of Total Cell Counts in BO Progressors and Non-Progressors E- Percentage distribution of Immune Cells in Barrett’s Oesophagus. CD4 cells demonstrate a more even percentage distribution between stroma and epithelium in patients that progressed from NDBO to OADC than non-progressors (stroma/epithelium percentage, progressor = 54.30/45.70 (p=0.370), non-progressor = 66.24/33.76 (p=0.001)).

**Table 3.**
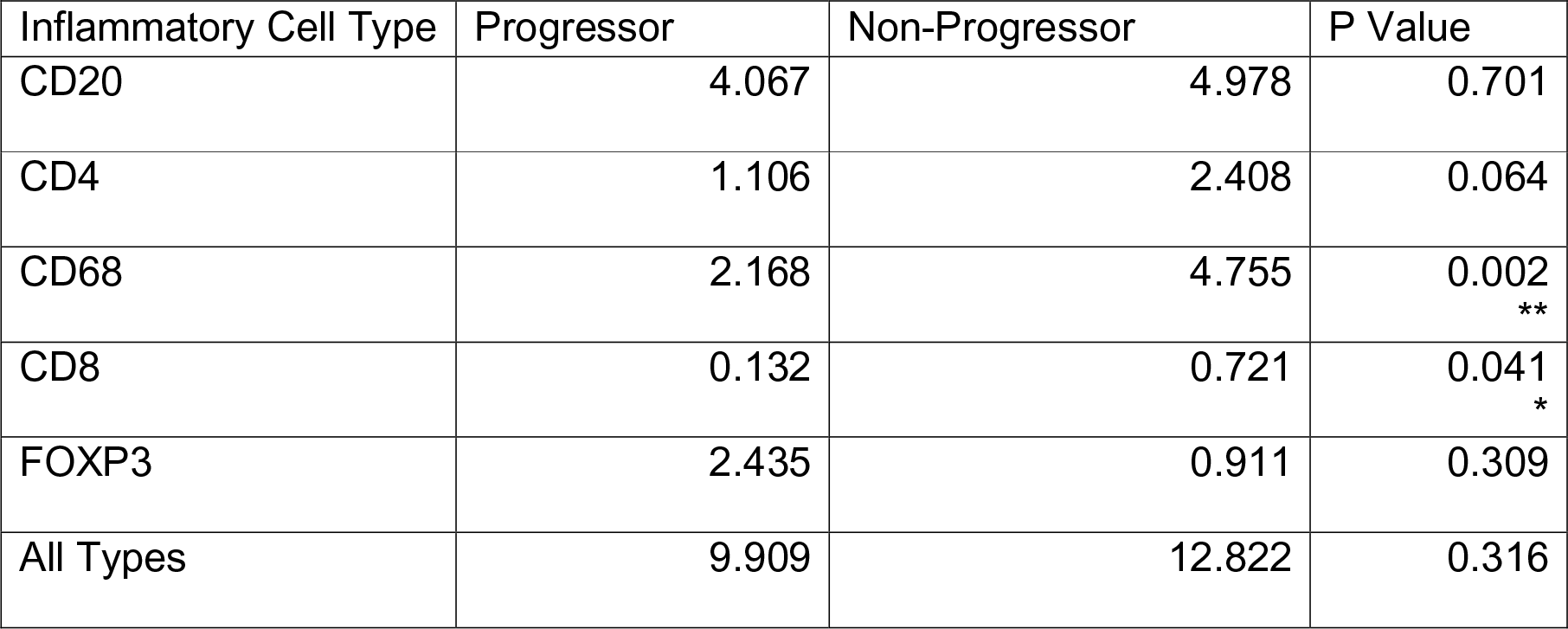
Inflammatory Infiltration of Immune Cells into BO Tissues. Average number of immune cells as a percentage of total cells. CD68 (p = 0.002, 95% CI = 1.134 to 4.038) and CD8 (p = 0.041, 95% CI - 0.02820 to 1.150) showed significantly higher infiltration in non-progressors then progressors. P-values * < 0.05 ** < 0.01

**Table 4.** Table showing the percentage of immune cell types in the stroma and epithelium of progressor vs non-progressor BO samples. There was no significant difference between progressors and non-progressors when stroma and epithelium were compared between the groups using an un-paired T-test.

| Inflammatory Cell Type | Non-progressor mean epithelial percentage | Progressor mean epithelial percentage | Non-progressor mean stromal percentage | Progressor mean stromal percentage | P Value |
| --- | --- | --- | --- | --- | --- |
| CD20 | 55.55 | 56.46 | 46.73 | 43.33 | 0.714 |
| CD4 | 35.11 | 45.70 | 66.24 | 54.30 | 0.294 |
| CD68 | 51.28 | 57.41 | 50.69 | 42.58 | 0.524 |
| CD8 | 22.02 | 23.55 | 80.73 | 76.45 | 0.850 |
| FOXP3 | 66.62 | 69.67 | 36.24 | 30.26 | 0.981 |
| Other | 69.29 | 75.34 | 31.66 | 24.58 |  |

**Table 5.** Average Percentage Distribution of Immune Cells in BO

|  | Progressor<br>Epithelial | Progressor<br>Stromal | p value | Non-Progressor<br>Epithelial | Non-Progressor<br>Stromal | p value |
| --- | --- | --- | --- | --- | --- | --- |
| CD20 | 56.46 | 43.33 | 0.123 | 53.09 | 46.73 | 0.523 |
| CD4 | 45.70 | 54.30 | 0.370 | 33.76 | 66.24 | 0.001<br>** |
| CD68 | 54.29 | 42.58 | 0.171 | 49.29 | 50.69 | 0.840 |
| CD8 | 17.05 | 66.28 | 0.001<br>** | 19.27 | 80.73 | <0.001<br>** |
| FOXP3 | 63.53 | 36.47 | 0.007<br>** | 63.76 | 36.24 | 0.010<br>* |

One non-progressor sample was excluded from the analysis as the total cell count was below 1000 (average total count = 20,300). Cell counts of each inflammatory cell type were compared to total cell counts in each tissue section to give a percentage value in progressor and non-progressor samples. We found significant decreases in CD68 positive cells (progressor = 2.168% vs non-progressor 4.755%. p = 0.002, 95% CI = 1.134 to 4.038) and CD8 (progressor = 0.132% vs non-progressor = 0.721%. p = 0.041, 95% CI - 0.02820 to 1.150), suggesting that progressor BO had reduced immune visibility. No further significant difference in immune cell infiltration was seen by individual type, and the overall inflammatory infiltrate when all subtypes were taken into account did not reach statistical significance (progressor = 9.909%, non-progressor 12.822%, p = 0.316, 95% CI = −4.011 to 11.74).

When differential immune cell distribution between stoma and epithelium was analysed together in each sample there was a non-significant difference between CD20 and CD68 cells which showed a relatively even distribution between stroma and epithelium in both groups. CD8 cells showed a stromal predominance whereas FOXP3 showed an increased epithelial distribution in both groups. When CD4 was compared, non-progressor samples showed a significant increase in stromal distribution of these cells (p=0.0014), whereas progressor samples showed that CD4 cells are more evenly distributed across the stroma and epithelium (p=0.37).

Together, we found a significant trend towards inflammatory infiltrate in non-progressive Barrett’s oesophagus, suggesting lack of immunovisibility. Overall, we conclude that the effect on OR3A4 over-expression seen in Barrett’s oesophagus is to drive an anti-inflammatory, MAPK signalling cascade that accelerates the progression towards dysplasia.

## DISCUSSION

In this study, we have investigated the role of the long non-coding OR3A4 in the pathogenesis of progression in Barrett’s oesophagus, the pre-malignant condition that can lead to invasive oesophageal adenocarcinoma.

We analysed transcriptional changes in CP-A cells with overexpressed OR3A4. The top two ranked differentially expressed pathways from RNAseq data showed two different possible mechanisms by which BO may progress to dysplasia and subsequent OADC – apoptosis and changes in MAPK signalling. We found that investigation of the human apoptosis pathway by RT-qPCR revealed upregulation of DIABLO, a gene (17) that encodes a protein that binds and inactivates ‘inhibitor of apoptosis proteins’ (IAPs)to promote caspase activation and apoptosis. This suggests that there is increased activation of apoptotic pathways in the OR3A4 over-expressing cells, possibly as a feedback mechanism in the absence of normally functioning apoptosis.

Additionally, we found that LT-α is downregulated in OR3A4 over-expressing cells. LT-α is a member of the tumour necrosis factor (TNF) family and is otherwise known as TNF-β. (18). LT-α forms a complex with LT-β, allowing binding to LT-β receptors (LTβR), promoting immune regulation through the innate immune response (19). This is especially interesting in the context of progression of BO to OADC as the loss of this pathway may contribute to changes in the immune recognition of the overexpressing BO cells (20). We observed that there was a decrease in TNF-β expression cells over-expressing OR3A4. If OR3A4 over-expression acts to inhibit the LTβR signalling mechanism it would result in reduced CD4 lymphocyte recruitment (21). In OR3A4 over-expressing cells, DIABLO is considerably upregulated, but without corresponding upregulation of TNF-β. The relative downregulation of LT-α and upregulation of DIABLO in the OR3A4 over-expressing may be a compensatory cellular mechanism to try to drive apoptosis in these cells.

We have additionally demonstrated increased phospho-p38 expression, suggesting that the p38 pathway is dysregulated. We have found an increase in expression of pERK, with fits with our observation of decreased DUSP4 (a negative regulator of ERK) (22) and increased p-p38 due to increased MAP2K3 expression in our RNAseq data. Downstream phospho-C-jun has reduced expression, suggesting dysfunction of the cell cycle due its role in increasing p53 signalling (23). We have also shown increased MAPK15 expression which is associated with increased migration and invasion in a c-Jun dependent manner, concordant with observations in colorectal cancer cell lines (24, 25) and in our wound scratch assays. We have also shown that Doramapimod, a p38 MAPK inhibitor which has been the subject of a number of Phase II clinical trials is active against OR3A4 over-expressing Barrett’s oesophagus cells, reducing p38 activation and decreasing viability (26). We did not find a similar change with aspirin therapy, suggesting that the mechanism of action of aspirin is not related to the p38 pathway in this context.

Consistent with the known role of p38 signalling in shaping the immune microenvironment (16), we observed, using multi-spectral IHC, that there was a significant decrease in the total numbers of CD8+ cytotoxic T cells and CD68+ macrophages in BO which progresses to OADC, suggesting that lack of immunovisibility may contribute to the progress of BO to OADC. This distribution may be tissue compartment specific, as we observed a change in the distribution of CD4+ T helper lymphocytes, with decreased stromal infiltration in BO progressing to OADC. Interestingly, Underwood et al have suggested that tumour stroma in OADC plays an important role in progression of OADC and pathological stroma subtypes can negatively affect patient survival (27). If CD8+ cells are not activated as readily due to a relative loss of CD4+ cells in the stroma, it may result in a proliferation of stromal CAFs. This may represent a further mechanism of progression to dysplasia and OADC.

Zingg et al (28) found that tumour infiltration of TREGs (FOXP3+) was not an independent prognostic factor in OADC. Similarly, Noble et al (29) did not find that Treg infiltration was independently prognostic in OADC or an independent marker for response to NAC. Reduction of the immune response to BO may be achieved by a number of mechanisms. In addition to the proposed mechanism here, a GWAS of patients with BO has demonstrated multiple predisposition SNPs in the Class I MHC region (30). This is important as the immune system relies on MHC class I (CD8+) and II (CD4+) for identification of foreign material for eradication.

The finding that OR3A4 which seems to be over-expressed in progressive BO drives a complex MAPK signalling cascade that is sensitive to dompiramod suggests future therapeutic pathways in targeting BO via this pathway may be possible.

## CONCLUSIONS

We have demonstrated that OR3A4 over-expression in Barrett’s Oesophagus is associated with an increase in p38 signalling and a reduction in immune infiltration that alters the immune microenvironment such that patients who progress to BO have a significantly reduced stromal immune infiltrate on the background of a proliferative epithelial phenotype. Further work is needed to understand the complex interplay between the p38 signalling and the immune system in Barrett’s Oesophagus.

## METHODS

### Cell lines

Cell culture was performed to analyse cell models and response to OR3A4 over-expression. HEK 293T, CP-A, OE-19, OE-21, OE-33 and MFD-1 cells were cultured during the period of study. All cells lines were incubated at 37°C with 5% injected CO2. The cell lines were split twice weekly at 90% confluence which provided an opportunity to refresh the media. All media was supplemented with 1 % penicillin/streptomycin antibiotic. Adherent cell lines were split by first removing the old culture medium and the confluent cell monolayer washed with sterile PBS to inactivate any remaining serum in the media. To ensure that only successfully transfected cells were represented in downstream experiments, un-transfected cells (CP-A, OE-19 and OE-33) were subjected to increasing doses of puromycin at 0, 0.25, 0.5, 1, 2 and 4 μg/ml in otherwise standard culture conditions.

The cell lines CP-A, OE-19, OE-33 and HEK-293T were transfected with a PIPZ lentiviral vector containing an OR3A4 over-expressing gene. This vector also contained a puromycin resistance gene to allow selection of successfully transfected cells. As a control, an empty vector was also transfected which still contained the puromycin resistance gene, but no OR3A4 over-expressing gene to ensure no adverse results were generated as a result of the transfection process. The Dharmacon pGIPZ lentiviral vector was modified by removing the green fluorescent protein reporter (GFP) and replacing it with a small multiple cloning site, into which the lncRNA OR3A4 was cloned. There was no short hairpin RNA (shRNA) cloned or expressed in the vector used.

HEK-293T cells are used as a packaging cell line so that the target gene sequence (OR3A4) can be integrated into the lentiviral plasmid using the host cell’s machinery. On day 1 HEK-293T cells were seeded at 2.5×10^5^ cells/ml into a 6 well plate. The following day lentivirus plasmids pMD2.G (150ng), psPAX2 (350ng) and gene of interest (OR3A4) (500ng), were transfected in the presence of Optimem media and Fugene transfection reagent. On day 3 the host cells (CP-A, OE-19, OE-21, OE-33 and HEK-293T) were seeded at 1×10^5^ cells/ml in a 6 well plate. The following day viral media was removed from transfected HEK-293T cells and mixed with an equal volume of with fresh host cell culture media, filtered through a PES 0.45μM filter, and placed on recipient cells.

On day 4, viral media was replaced with fresh host cell culture media containing 1μg/ml puromycin. Control (non-infected) cell death was used as a marker of successful transduction.

### Immunofluorescence

Cell morphology was assessed using light microscopy and IF. Low and high power views were obtained of control and OR3A4 over-expressing cell lines CP-A, OE-19 and OE-33 and digital photographs taken using the EVOS FL Cell Imaging System (ThermoFisher). For IF images, cells were cultured on slides, fixed and stained with phalloidin, DAPI counter nuclear stain and cleaved caspase 3 IF images were obtained using a Vectra confocal microscope. For immunofluorescence, Phalloidin, Caspase 3 and Hoescht (nuclear counter stain) stains were used. An indirect staining method with primary and secondary antibodies was performed. To prepare the slides, cells were grown in a monolayer on sterile coverslips in media. Once the cells had adhered and reached the desired levels of confluence, the coverslips were removed from the media and placed in a clean dish for the staining process. The cells were washed twice with PBS at 37C taking care not to dislodge the adherent cells from the coverslip. The cells were then fixed with 4% Paraformaldehyde in PBS for 10 minutes. The cells were then washed with PBS 3 times for 5 minutes. To allow the stain to penetrate the cells, the membrane was permeabilised with 0.1% Triton diluted in PBS for 2 minutes. The cells were again washed with PBS 3 times for 5 minutes. Non-specific antibody binding was blocked by adding a solution of 10% heat inactivated goat serum for 40 minutes at room temperature.

Rabbit primary monoclonal antibody was diluted to a 1:400 with PBS (6 μL antibody to 2.4 mL PBS). Parafilm was stretched over the lid of a 6 well culture plate. A 200 μL globule of the antibody solution was pipetted on to the parafilm. The coverslip was carefully transferred on to the antibody solution with the cells in contact with the antibody. This was transferred to the fridge for overnight incubation. The coverslip was washed three times with PBS to remove the primary antibody solution. The secondary antibodies were prepared with a 1:400 dilution (4 mL PBS, 10 μL secondary anti-rabbit, 100 μL phalloidin/caspase 3). Cell Signalling Technologies Cleaved Caspase 3 (ASP175) and Alexa Fluor 594 Phalloidin (Invitrogen) were used, along with secondary Goat Anti-rabbit Alexa Fluor 488 (Invitrogen). 300 μL of the secondary antibody solution was placed on each coverslip and incubated for 40 minutes at room temperature. Following the incubation the coverslips were washed three times with PBS. Hoechst DNA counter stain was prepared with a 1:1000 dilution and applied to the coverslips for 5 minutes. The coverslips were then mounted on slides and allowed to air dry. Once dry, they were sealed with cytoseal. The slides were then imaged using the Vectra confocal microscope, and image processing for size measurements was carried out using QuPath v.0.2.0-m4.

### Cell viability/proliferation assays

To assess the effect of OR3A4 expression on cell viability and growth rates, MTS and scratch wound healing assays were performed. For MTS assay for cell viability, 10^3^ Cells were seeded into each well of a clear bottomed 96 well culture plate and media added to make up the total volume in each well to 100μL. Blank wells containing media alone were also included on the plates to allow background absorbance to be corrected. The plates were incubated for 24 hours to allow the cells to adhere to the culture plate. Enough wells were seeded to allow MTS assays to be run over the course of 5 days. At 24, 48, 72 and 96 hours post seeding, 20μL of the CellTitre96 MTS reagent was added to each well to be studied including the media blanks. The plate was returned to the incubator and read on the plate reader at 1, 2, 3 and 4 hours after adding MTS reagent. The absorbance of the control blank wells was subtracted from the absorbance of the experimental wells to give a change in absorbance.

Ibidi culture plates with 2 well inserts were used for scratch wound healing assays. Specialised culture plates with inserts were used following many aborted attempts to generate a regular scratch wound in a cultured monolayer of cells by hand. Adherent cells were trypsinised when they reached 90% confluence and re-suspended in culture media. 70 μL of the cell suspension was pipetted into each of the wells on the plate insert. Once the cells had re-attached and reached 100% confluence, the plate inserts were removed to create a 500 μm cell free gap. The culture plates were imaged at regular time intervals until the cell free gap was completely closed. The cell free gap was measured using ImageJ software.

### Drug screening

The cell lines CP-A, CP-A PIPZ control and CP-A PIPZ OR3A4 were selected for drug treatment with the p38 MAPK inhibitor Doramapimod (Selleckchem S1574) and Aspirin (Acetylsalicylic acid, Sigma-Aldrich A2093). In order to assess their viability following drug treatment they were subjected to MTS assays. 1000 cells were seeded in each well of a white walled 96 well plate containing 100 μL media. After 24 hours each well was dosed with either aspirin, doramapimod or a combination of agents with increasing concentrations. Aspirin was dosed from 0-100 mM concentration. Doramapimod was dosed initially from 0-10 nM, then subsequently from 0-10 μM. This was taken as day 1, and an immediate MTS assay performed. MTS reagent was added after a further 24, 48, 72 and 96 hours and the plate re-read.

### RNA analysis

Total RNA from CPA, CPA PIPZ Control, CPA PIPZ OR3A4, OE33, OE33 PIPZ Control and OE33 PIPZ OR3A4 cells was sequenced using the Illumina NextSeq 500 platform. RNA was extracted using the Qiagen RNeasy kit and QC’d using the Qubit fluorometer and Agilent Tapestation 2200. It was then subject to automated library preparation using the Illumina NeoPrep instrument and following the TruSeq Stranded mRNA Library Prep for NeoPrep protocol. Samples were sequenced to an average of 25M paired-end reads.

For real time quantitative polymerase chain reaction (RTqPCR), RNA was extracted using the Qiagen RNeasy kit as described above. This was then converted to cDNA with the Applied Biosystems High Capacity RNA to cDNA Kit (4387406). RTqPCR was performed on cDNA generated from RNA extracted from CP-A, CP-A PIPZ Control, CP-A PIPZ OR3A4, OE-33, OE-33 PIPZ Control, OE-33 PIPZ OR3A4, OE-19 and MFD-1 cell lines in the manner previously described in methods sections 4.5 and 4.16. This was analysed on the Quantstudio 7 Flex Real-Time PCR System (ThermoFisher).

CP-A, CP-A PIPZ control and CP-A PIPZ OR3A4 cell lines were treated with the p38 MAPK inhibitor Doramapimod and Aspirin in increasing doses and MTS assays performed daily up to 5 days incubation to assess cell viability.

### Multi-parametric quantitative immunohistochemistry

Ethical approval for retrieval of pathology specimens from the histopathology archive was obtained from the NW Research Ethics committee (ref 15/NW/0079) using anonymised biobanking consent. A panel of CD4, CD8, CC20, CD68 and FOXP3 was used for multispectral IHC. All sections were reported as non-dysplastic on histological review (Supplementary table 1). The manufacturers recommended dilution was used on each antibody and a two-fold increased and decreased dilution was used to find the optimal condition with DAB staining. The optimal dilutions are detailed in

Supplementary table. The optimal dilution for each antibody was paired with each of the opal dyes (OPAL 520, OPAL 540, OPAL 570, OPAL 620, OPAL 650 and OPAL 690), to find out which dye was compatible. Single antibody/opal dye combinations were used for the spectral library and the best combination of antibody/opal dye was combined in sequential staining. The order of antibodies was switched to find the optimal panel combination. The optimal panel combination was achieved when the multiplex image was comparable to that of the single IHC. Once the optimal antibody dilutions and conditions were finalised, the FFPE sections were heated to 60C to de-wax. The stains were added in the sequence shown in

Supplementary table. The sections were re-hydrated and subjected to heat induced epitope retrieval (HIER). The CD4 antibody underwent pre-staining HEIR in BOND Epitope Retrieval Solution 2, the other antibodies used BOND Epitope Retrieval Solution 1. The sections were heated to 100C for 20 minutes in the Epitope Retrieval Solution. Following HEIR, the primary antibody was added. Secondary rabbit, anti-mouse antibodies along with the corresponding Tyramide Signal Amplification (TSA) OPAL dye were then added. The sections were stained in the sequence: CD4, CD8, CD20, FOXP3, CD68. Finally DAPI nuclear counter stain was used.

The VECTRA 3 automated quantitative pathology imaging system was used to scan the slides. To create a spectral library, single stained slides, OPAL 520, OPAL 570, OPAL 620, OPAL 650, OPAL 690, DAPI and an auto fluorescence slide were scanned. All filters were used at x20 magnification. A spectral library was created in the InForm software from the images. For multiplex scanning, whole slides were scanned at x10 magnification and at x20 for multispectral imaging. Each multiplex slide was exposed with each of the five available filters: DAPI, FITC, CY3, Texas Red and CY5, ensuring the slide was in focus with each of the images.

The multispectral IHC images were analysed using the Perkin Elmer inForm Cell Analysis Software. This is a software package with advanced image processing capabilities which utilises a machine learning algorithm to recognise different cell and tissue types within a histopathology slide image. Once the images are loaded on to the software, the different cell types are manually selected by the user. At least 8 of each differentially stained cell type must be identified for the algorithm to accurately detect them in new, previously “unseen” images. Once the cell types are identified, different zones within the tissue section are marked. Background, stroma and epithelium is manually traced by the user using the computer mouse. The software algorithm processes the information and attempts to train and then delineate the different zones on unseen images. Although the tissue sections were all from BO, stained in a homogenous manner, the algorithm required training on numerous different sections in order to begin automated recognition of tissue zones.

Once the software reliably de-lineated epithelium and stroma, the finalised algorithm was run with all sections of all samples to generate a count of immune cells and their distribution across the tissue sections. Microsoft Excel pivot tables were used to collate the data and determine the distribution of cells.

## Data Availability

All data available on request.

## Experimental design

TN, MC, ADB

## Experimental work

TN, YS, QQZ, JS, CW, RH, AS, VP

## Manuscript

All authors

## Statistical analysis

Andrew Beggs

## Data sharing statement

All sequence data will be uploaded to the SRA archive on publication of this study. All methylation microarray data will be uploaded to the GEO archive on publication of this study.

---

**Supplementary table 1.** Antibodies used for multispectral IHC including staining time, optimal dilutions, TSA OPAL marker dyes and secondary antibodies. The antibodies were stained in sequence one through five.

| Antibody | Stain Order | Clone | Ab time | Ab dilution | TSA OPAL | TSA dilution | TSA time | PRE-TREAT | Secondary |
| --- | --- | --- | --- | --- | --- | --- | --- | --- | --- |
| **CD4** | 1 | 4B12 | 30 | 1:50 | 520 | 1:400 | 10 | Ph9 20 | mouse 1:100<br>(dako RABBIT ANTI MOUSE) |
| **CD8** | 2 | 4B11 | 30 | 1:400 | 570 | 1:400 | 10 | pH6 10 | mouse 1:100<br>(dako RABBIT ANTI MOUSE) |
| **CD20** | 3 | L26 | 30 | 1:200 | 620 | 1:200 | 10 | pH6 10 | mouse 1:100<br>(dako RABBIT ANTI MOUSE) |
| **FOXP3** | 4 | 236A/<br>E7 | 30 | 1:400 | 650 | 1:400 | 10 | pH6 10 | mouse 1:100<br>(dako RABBIT ANTI MOUSE) |
| **CD68** | 5 | KDI | 30 | 1:400 | 690 | 1:200 | 10 | pH6 10 | mouse 1:100<br>(dako RABBIT ANTI MOUSE) |

